# Natural Mucosal Barriers and COVID-19 in Children

**DOI:** 10.1101/2021.02.12.21251310

**Authors:** Carl A. Pierce, Sharlene Sy, Benjamin Galen, Doctor Y Goldstein, Erika Orner, Marla J. Keller, Kevan C. Herold, Betsy C. Herold

**Author notes:** Contributed equally (Kevan C. Herold, Betsy C. Herold).

## Abstract

COVID-19 is more benign in children compared to adults for unknown reasons. This contrasts with other respiratory viruses where disease manifestations are often more severe in children. We hypothesized that a more robust early innate immune response to SARS-CoV-2 may protect against severe disease and compared clinical outcomes, viral copies and cellular gene and protein expression in nasopharyngeal swabs from 12 children and 27 adults upon presentation to the Emergency Department. SARS-CoV-2 copies were similar, but compared to adults, children displayed higher expression of genes associated with interferon signaling, NLRP3 inflammasome, and other innate pathways. Higher levels of IFN-α2, IFN-γ, IP-10, IL-8, and IL-1β protein were detected in nasal fluid in children versus adults. Anti-SARS-CoV-2 IgA and IgG were detected in nasal fluid from both groups and correlated negatively with mucosal IL-18. These findings suggest that a vigorous mucosal immune response in children compared to adults contributes to favorable clinical outcomes.

## Introduction

Epidemiological studies have consistently shown that children infected with severe acute respiratory syndrome coronavirus 2 (SARS-CoV-2) have a milder clinical course with significantly less morbidity and mortality than adults. The Centers for Disease Control (CDC) estimates that approximately 1.2-3.3% of total hospitalizations and less than 0.21% of deaths from coronavirus disease 2019 (COVID-19) are in children (1). This experience is in contrast to other respiratory viruses, such as influenza or respiratory syncytial virus, where disease manifestations in children are often more severe than adults (2). Several hypotheses have been proposed to explain why children are protected from more severe outcomes with COVID-19 including differences in expression of angiotensin-converting enzyme 2 (ACE2), the receptor for viral entry, resulting in lower viral loads, presence of antibodies to common cold coronaviruses that might provide partial protection, and a more robust innate response early in the course of infection that mitigates against a vigorous adaptive response (3, 4). However, recent studies have shown that ACE2 expression is not reduced in children and may actually be lower in adults (5). Surveys of children infected with COVID-19 have found that the amount of SARS-CoV-2 RNA detected in nasopharyngeal (NP) swabs is at least as high in children compared to adults (6). It is also unlikely that antibodies that are cross reactive to other viruses explain the clinical differences as we previously found that antibody levels to other common cold human coronaviruses (229E, NL63, HKU1) were similar in adults and children. In addition, although common cold coronavirus antibody levels may be boosted in response to SARS-CoV-2 infection, they do not provide protection (7, 8).

In a previous analysis of patients hospitalized with COVID-19, we found that serum levels of IL-17A and IFNγ collected early in the course of disease were higher in children versus adults and correlated significantly and inversely with age (7). The source of these cytokines did not appear to be peripheral blood mononuclear cells as adults had more robust T cell responses. These findings suggested that other cells, such as innate immune cells within the respiratory tract, might be the source of the IL-17A and IFN-γ and that a more vigorous mucosal innate response could account for the milder clinical course in children. To test the hypothesis that age-related outcomes reflect differences in host responses at the site of viral exposure, we collected nasopharyngeal (NP) swabs from pediatric and adult patients who presented to the Emergency Department at Montefiore Medical Center in The Bronx, NY, analyzed the harvested cells for gene expression, and measured the levels of cytokines and antibodies in the mucosa.

## Results and Discussion

The demographics and clinical characteristics of the study population are described in Table S1. There were no significant differences in number of days of symptoms prior to presentation or the levels of SARS-CoV-2 viral RNA (quantified by cycle threshold [Ct] values) in NP swabs comparing the pediatric (n=12) and adult (n=27) patients (Fig. 1A). However, the outcomes differed. Adults were more likely to be admitted to the hospital (81% vs 42%, p=0.02), had higher C-reactive protein (10.48 ± 7.19 vs 6.03 ±10.48 mg/dl, p<0.0001) and D-dimer (2.38 ± 3.42 vs 0.81 ± 0.59 μg/ml, p<0.0001) at admission, and if hospitalized, had a longer length of stay (10.36 ± 10.85 vs 3.0 ± 1.73 days, p<0.0001) (Table S1). None of the pediatric patients required oxygen whereas 7 adults did and 4 required mechanical ventilation (p=0.03). Four adults but none of the pediatric patients died.

**Figure 1:**
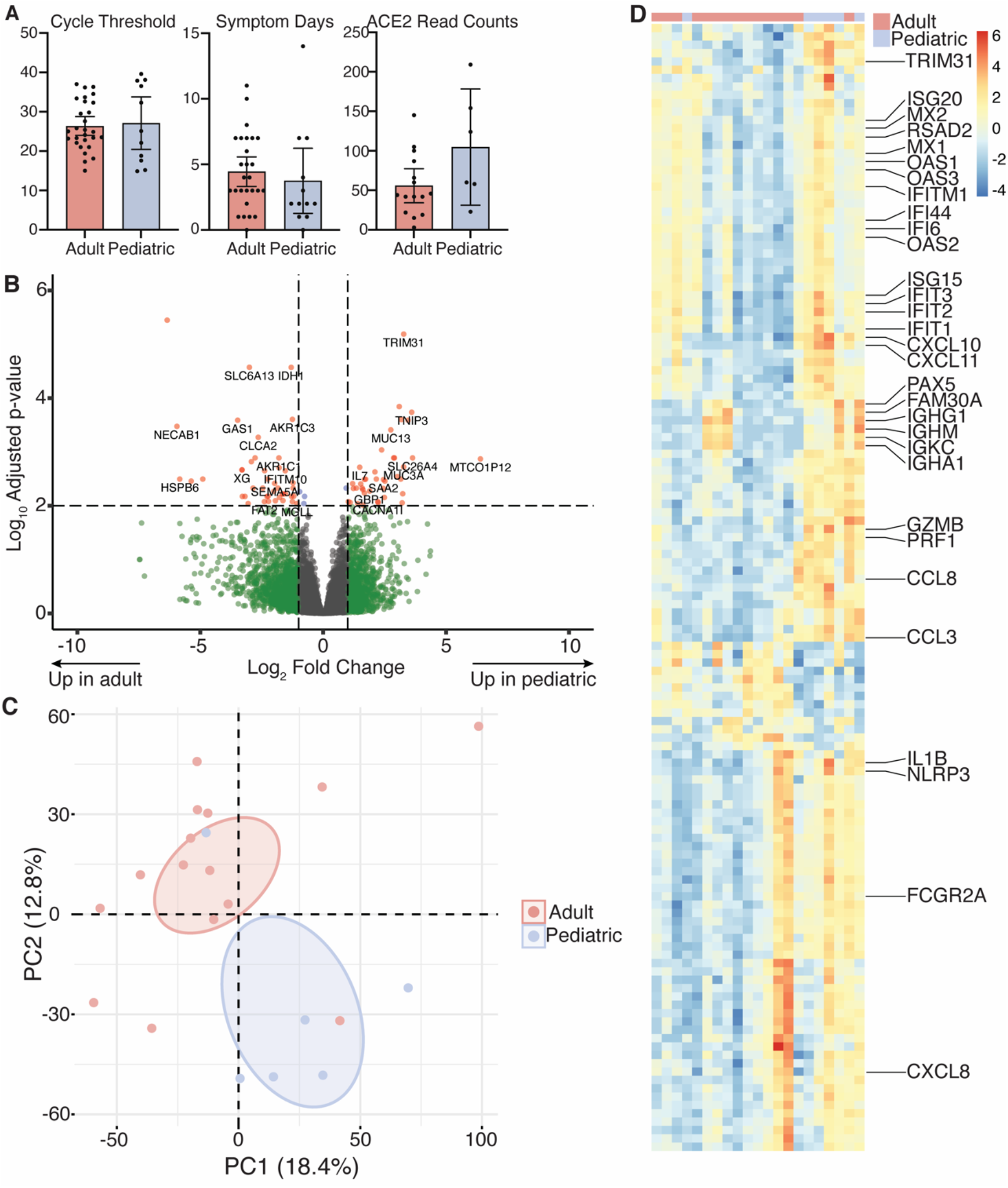
Transcriptional profiles in pediatric and adult nasopharyngeal samples. (**A**) Cycle threshold (Ct) values and days of symptoms prior to presentation in 27 adult and 11 pediatric patients, and ACE2 read counts in cells isolated from NP swabs obtained at presentation in 15 adult and 6 pediatric samples. Bars show mean ± 95% CI. (**B**) Volcano plot of genes expressed more strongly in pediatric (right) and adult (left) patients. (**C**) Principal component plot of RNAseq data; n = 15 adult and 6 pediatric samples. Ovals are 95% confidence ellipses. (**D**) Heatmap showing expression of the top fifty contributing genes in principal components one, two, and three.

To evaluate whether the age-related differences in outcomes reflect differences in host responses at the site of viral exposure, the nasal mucosa, we performed RNA-Seq on cells isolated from NP swabs obtained at the time of presentation to the Emergency Department (n=6 pediatric and 15 adult patients). We identified 538 differentially expressed genes (p<0.05 after FDR correction) including 267 that were upregulated and 271 that were downregulated comparing pediatric and adult patients (Fig. 1B). Notably, expression of ACE2 trended towards being higher in the pediatric samples (p=0.053, Fig 1A).

Principal component analysis (PCA) of the RNAseq dataset separated pediatric and adult patients into non-overlapping clusters with the first dimension capturing 18.4% of the overall variance, the second dimension capturing 12.9% and the third 7.9% (Fig. 1C). To understand the gene expression patterns contributing to the segregation of the samples, we interrogated the 50 genes contributing most strongly to the principal components (Table S2). This gene set was enriched for interferon-stimulated and other innate immune response genes such as IL-1β, CCL3, CXCL10, and NLRP3 (Fig. 1D). After dichotomization of adults by clinical outcome, PCA analysis showed that adults who never required supplemental oxygen were intermediate between adults who required respiratory support and children (Figure S1).

Given the strong contribution of innate immune gene expression in our PCA, we performed Gene Set Enrichment Analysis to evaluate the immune landscape. This showed enrichment of genes in the IFN-γ response (hallmark:M5913, Normalized Enrichment Score (NES)=3.66, p=0.006), IFN-α response (hallmark:M5911, NES=3.52, p=0.006), IL-1 response (GO:0070555, NES=2.70, p=0.03), NLRP3 inflammasome (GO:0072559, NES=1.88, p=0.03), and IL-17 production (GO:0032620, NES=2.30, p=0.03) in pediatric relative to adult patients (Fig. 2A). Conversely, fatty acid metabolism (hallmark:M5935, NES=-1.86, p=0.006) was enriched in adults compared to pediatric patients (Fig. 2B). Oxidative phosphorylation and glycolysis pathways were also increased in adults versus children, but these were not statistically different after FDR correction (NES=-1.18, p=0.14; NES=-1.09, p=0.3, respectively). These findings further support an enhanced innate response to SARS-CoV-2 infection in children but enhanced metabolic pathways among cells from adults.

**Figure 2:**
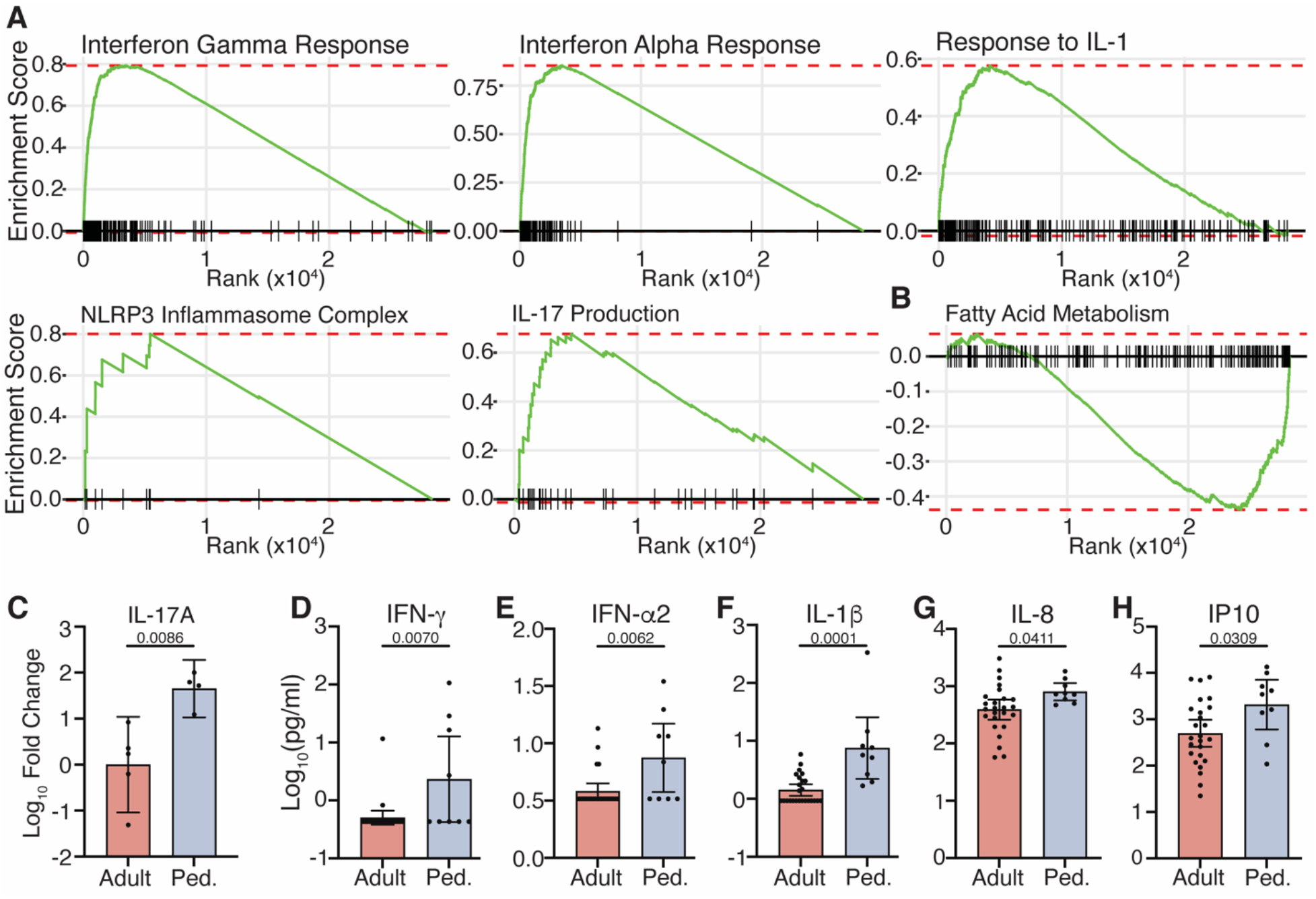
Innate responses in pediatric and adult nasopharynx. (**A**) Gene set enrichment plots for the indicated pathways: IFN-γ response (Normalized Enrichment Score (NES)=3.66, p=0.006), IFN-α response (NES=3.52, p=0.006), IL-1 response (NES=2.70, p=0.03), NLRP3 inflammasome (NES=1.88, p=0.03), and IL-17 production (NES=2.30, p=0.03). (**B**) Fatty acid metabolism (NES=-1.86, p=0.006). All plots show enrichment in RNAseq data from pediatric patients relative to adult patients. **(C)** Relative IL-17A gene expression measured by RT-qPCR in 5 adult and 4 pediatric samples not used for RNAseq. Fold change was calculated by the 2^CT(ref)-CT^ method using the mean adult value as the reference. **(D-H)** Levels of the indicated cytokines in NP transport media from 25 adult and 9 pediatric patients measured by multiplex Luminex assay. p-values are listed above comparison bars. Unpaired t-test (**G, H**) or Mann Whitney test (**C-F**). Ped = pediatric. Bars show mean ± 95% CI.

We then performed RT-qPCR using RNA isolated from NP swabs from an additional 4 pediatric and 5 adult patients who were not included in the RNA-Seq analysis to confirm these findings. IL-17A gene expression was significantly increased in children (Fig. 2C) and there were similar trends for higher levels of IFN response genes (Figure S2). We also measured cytokine levels in NP fluid and found significantly higher levels of IFN-γ, IFN-α2, and IL-1β, IL-8 and IP10 in pediatric patients (Fig. 2D-H), consistent with the RNAseq data.

SARS-CoV-2-specific IgA in nasal mucosal secretions may contribute to protection (9) and our RNA-Seq data revealed a cluster of B cell-associated genes in the PCA gene set (Dim 2; *IGHA1, IGKC, IGHM, IGHG1, FAM30A and PAX5*) with higher expression in a subset of both adult and pediatric samples (Fig 1D). Therefore, we measured total IgA and IgG by ELISA and SARS-CoV-2 specific IgA and IgG targeting S1, S2, and the receptor binding domain (RBD) of the spike and nucleocapsid (NC) protein by multiplex assay in the NP fluid. We did not identify differences in total IgG or IgA comparing pediatric and adult patients (Fig. 3A, C). While COVID-19 patients had elevated levels of SARS-CoV-2-specific immunoglobulins compared to healthy controls (n=7 adults), there were no significant differences between age groups (Fig. 3B, D). Agglomerative hierarchical clustering using only the B cell-associated genes dichotomized the cohort into a “low” or a “high” expression group (Fig. 3E). All of the adult patients with more severe clinical disease who required supplemental oxygen (SO) were in the low transcript group (Fig. 3E).

**Figure 3:**
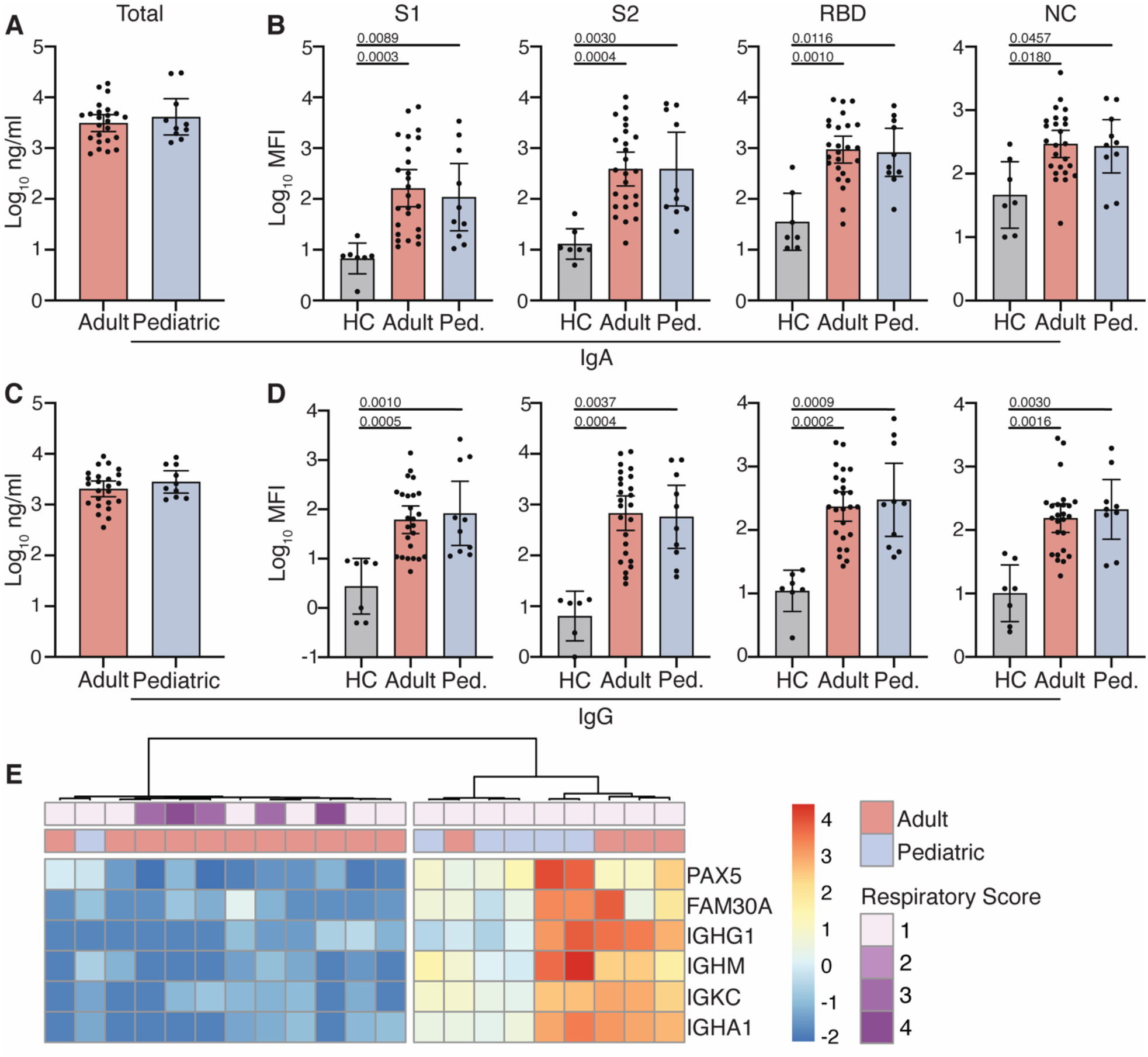
Early mucosal antibody responses in pediatric and adult COVID-19 patients. **(A)** Total and (**B**) SARS-CoV-2 specific IgA and **(C)** total and (**D**) SARS-CoV-2specific IgG levels at time of presentation were measured in 10 pediatric and 25 adult patients and 7 healthy controls (HC). (**E**) Heatmap showing expression of B cell-related genes contributing to PC1-3. Annotations show age group and peak respiratory score (1 = Room Air, 2-4 = Supplemental Oxygen). Total antibody levels (**A, C**) measured by ELISA; SARS-CoV-2 antibody levels (**B, D**) measured by multiplexed Luminex assay. Where significant, p-values are listed above comparison bars; Kruskall-Wallis (**B, D**). Ped. = pediatric. Bars show mean ± 95% CI.

Correlation matrices were generated to explore possible associations between NP antibody levels, cytokines, and SARS-CoV-2 Ct values (Fig. 4A). In general, antibodies and cytokines did not strongly correlate with one another. However, we observed a significant and inverse relationship between SARS-CoV-2-specific antibodies (IgG and IgA) and IL-18 (Fig. 4C, D). There were no differences in IL-18 levels comparing children and adults, but there was a trend towards lower IL-18 in adults who required supplemental oxygen (Fig. 4B, p=0.12).

**Figure 4:**
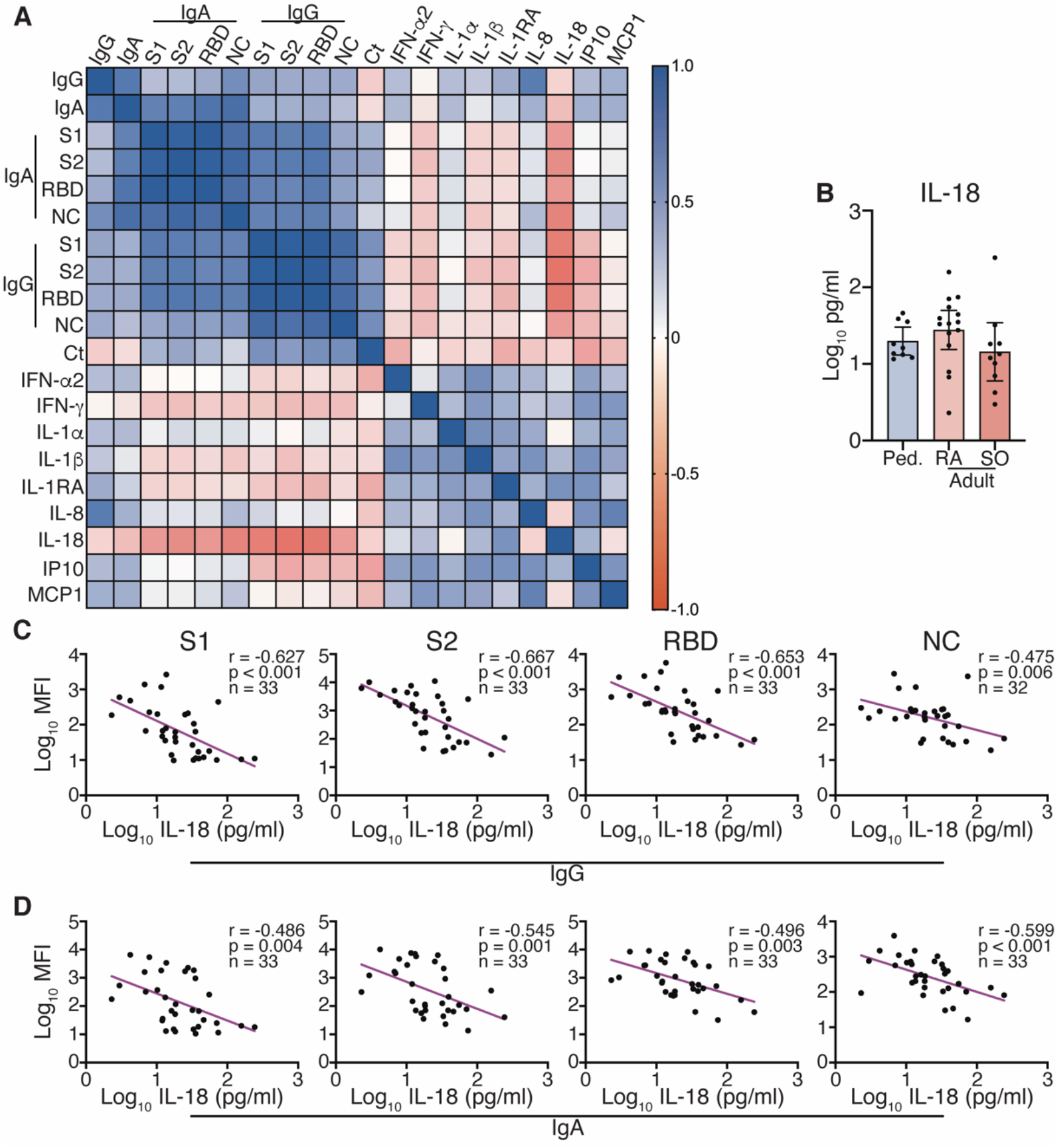
Nasopharyngeal IL-18 levels negatively correlate with SARS-CoV-2-specific mucosal antibody. **(A)** Spearman correlation matrix of Ct values, antibody, and cytokine data. **(B)** IL-18 levels in pediatric patients and adults who did not (RA) or did (SO) require supplemental oxygen. **(C)** Correlations between IL-18 and SARS-CoV-2-specific IgG. **(D)** Correlations between IL-18 and SARS-CoV-2-specific IgA. Ped = Pediatric (n=9); RA = Room Air (n=15); SO = Supplemental Oxygen (n=10). Bars show mean ± 95% CI

Our studies identify age-related differences in primary immune responses to SARS-CoV-2 at the nasal mucosa, the presumptive site of first viral encounter, which may contribute to the clinical outcomes. Most studies focus on systemic immune responses and have measured cytokines and antibodies in the blood, but our prior and other studies led us to speculate that the innate immune response was more vigorous in children (7). Our new findings using NP swabs obtained at the earliest time point of presentation establish this and suggest that a more robust mucosal innate response in children overcomes viral evasion strategies and generates an immediate barrier to viral infection. This may dampen the subsequent adaptive immune response as evidenced by our prior findings of lower neutralizing antibodies, decreased antibody-dependent cell mediated phagocytic activity (ADCP), and less robust T cell responses in children versus adults (7). A reduced adaptive response in children compared to adults has been confirmed by others who documented decreased neutralizing antibody titers (10) and less robust T cell responses (11).

In contrast, a reduced or delayed mucosal response in adults, evidenced by the decreased expression of transcripts associated with innate pathways in their NP swabs may lead to an inability to escape viral immune evasion and a more vigorous adaptive response (12, 13). The latter contributes to high systemic levels of other inflammatory cytokines (e. g. IL-6 and TNF) and an increased risk of acute respiratory distress syndrome, which has also been observed with SARS-CoV-1 (14–17). Other studies have found an impaired innate response to SARS-CoV-2 in adults compared to other respiratory viral infections (18).

Children have more frequent respiratory infections than adults. This, as well as recent childhood immunizations, could contribute to a higher basal level of activation of innate responses as suggested by studies of rhinovirus in which an early interferon response was associated with rapid viral clearance (19). Notably, a recent study found that infection of organoid cultures with rhinovirus protected against subsequent SARS-CoV-2 challenge (20). The prior respiratory infections in children may prime innate immune cells that can rapidly respond to virus. The higher gene expression of metabolic flux pathways in adults may reflect the metabolic demands needed to activate innate pathways (21) and may account for a delayed response to COVID-19 (22).

We detected similar levels of SARS-CoV-2 specific IgA and IgG in NP samples obtained from adults and children at this early time point before a fully mature antibody response would be expected. Notably these early antibody responses correlated inversely with mucosal IL-18 levels.

IL-18, an IL-1 superfamily cytokine predominantly produced by macrophages, is cleaved to its active form by the NLRP3 inflammasome and ultimately promotes the production of IFN-γ. We speculate that the early release of this cytokine may temper the adaptive response which is consistent with the more severe outcomes with lower levels of IL-18. The kinetics of its secretion may be important since at later time points, elevated serum IL-18 levels are associated with increased inflammasome activation and disease severity (23).

In summary, we show, for the first time, direct evidence for a more vigorous early innate immune response in the nasopharynx of children compared to adults at clinical presentation with COVID-19. Innate immune responses to other pathogens have also been shown to decrease with age (24, 25) The cellular source of these protective cytokines is not clear but could include mucosal and airway epithelial or invariant natural killer T cells (26, 27). Nonetheless, our findings suggest that airway resident cells establish the response to virus that ultimately determines the clinical outcomes. The reasons for the differences in the early innate responses with age are not clear, but therapies that enhance these pathways may be an effective form of treatment and protect from further damage from viral invasion as suggested by ongoing trials of inhaled interferon beta-1 for early treatment of COVID-19 (28).

## Methods

### Study design

NP swabs were obtained from the Montefiore Clinical Laboratory from patients with confirmed SARS-CoV-2 infection by PCR assay who presented to the Emergency Department at Montefiore Medical Center between November 2020 and January 2021. Patients were excluded if they had pre-existing medical conditions that might impact immune responses including cancer and HIV, pregnancy, or were receiving chronic immunosuppressive therapy. Demographics, length of stay, peak respiratory support required (1=room air, 2=nasal cannula; 3=CPAP or high flow nasal oxygen, and 4=mechanical ventilation), outcome and the results of clinical laboratory studies including SARS-CoV-2 PCR (Ct values), complete blood counts, D-dimer, and C-reactive protein were obtained by chart review.

The NP swabs were collected in viral transport medium. A portion was removed for measurement of SARS-CoV-2 RNA by PCR in the Clinical Microbiology Lab and the remainder of the sample was transported. NP swabs were also obtained from 7 healthy controls. The swab was transferred to a tube containing media supplemented with 13 µM dithiotrietol to inactivate virus and then incubated in a thermomixer for 10 minutes at 37°C and 500 RPM. The cells were washed and frozen in 500µl 90% FBS/10% DMSO. The original transport medium (nasal fluid) was aliquoted and stored at −80°C for measurement of cytokines and antibodies.

### RNA sequencing

RNA was isolated from cryopreserved cells using the miRNEAsy Micro kit (Qiagen 217084). Samples with sufficient RNA quantity and quality were used for analysis. The methods for library preparation, sequencing, and analysis are discussed in Supplementary Methods. For quantitative RT-PCR, RNA was isolated as for RNAseq and cDNA synthesis was performed with methods described in Supplementary Methods.

### Cytokine and Antibody Measurements

Media from NP swabs was thawed and treated with UV light to inactivate virus prior to use. SARS-CoV-2-specific IgG and IgA were measured using MILLIPLEX SARS-CoV-2 Antigen Panel 1 IgG and IgA kits (Millipore HC19SERG1-85K and HC19SERA1-85K, respectively). Cytokines were measured using the MILLIPLEX Human Cytokine/Chemokine/Growth Factor Panel A kit (Millipore, HCYTA-60K).

### Statistical Analyses

Because of sample volume limitations not all assays could be performed with all samples. Missing data was at random and the number of samples used are indicated. Statistical analyses were performed in GraphPad Prism (v9.0.1, GraphPad Software, Inc). Cytokine and antibody data were log transformed prior to analysis. Normality was tested and a parametric or non-parametric test was used for comparison of groups as indicated.

### Study Approval

This study was approved by the Institutional Review Board of the Albert Einstein College of Medicine (IRB# 2020-11278). Written informed consent was obtained for samples from healthy controls.

## Data Availability

Data will be deposited in a public database and accession codes provided at time of publication.

## Author Contributions

Study design: CAP, KCH, BCH; Performance of experiments: CAP; Data acquisition and analysis: BG, SS, DYG, EO, MJK; Writing and editing: CAP, MJK, BCH, KCH.

## Acknowledgments

Supported by NIH grants R01 AI134367 (BCH and KCH), UL1 TR002556, P30 AI124414, an Einstein College of Medicine Dean’s COVID-19 Pilot Research Award (BH). CAP is supported by T32 AI007501, NIGMS MSTP training grant T32GM007288 and The Eric J. Heyer, MD, PhD Translational Research Pilot Project Award.

---

**Figure S1:**
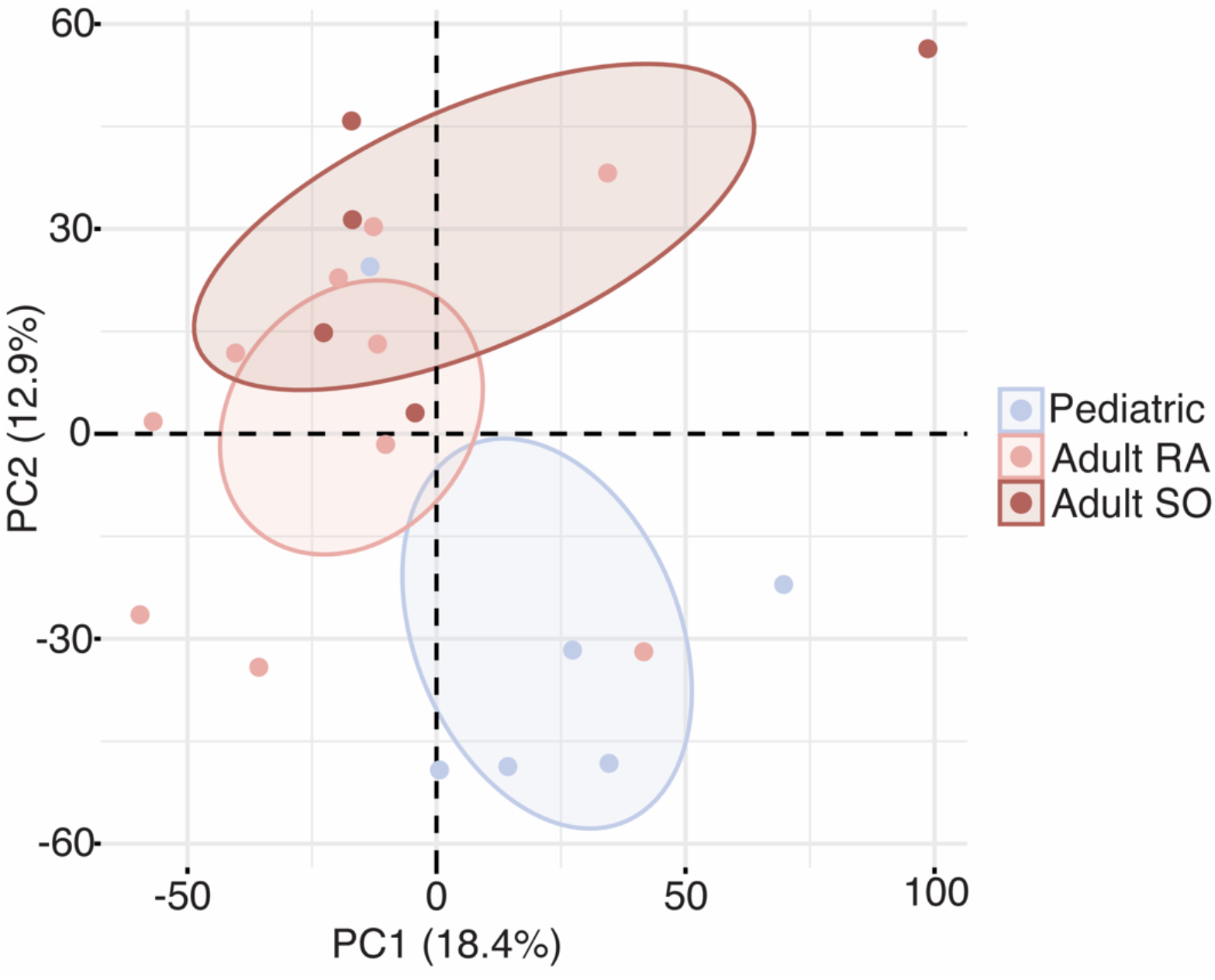
Adults not requiring oxygen are intermediate between children and adults who did. Principal component plot showing 95% confidence ellipses for pediatric samples, and samples from adults who did or did not require supplemental oxygen. RA = Room Air; SO = Supplemental Oxygen. N = 6 pediatric, 10 RA, 5 SO.

**Figure S2:**
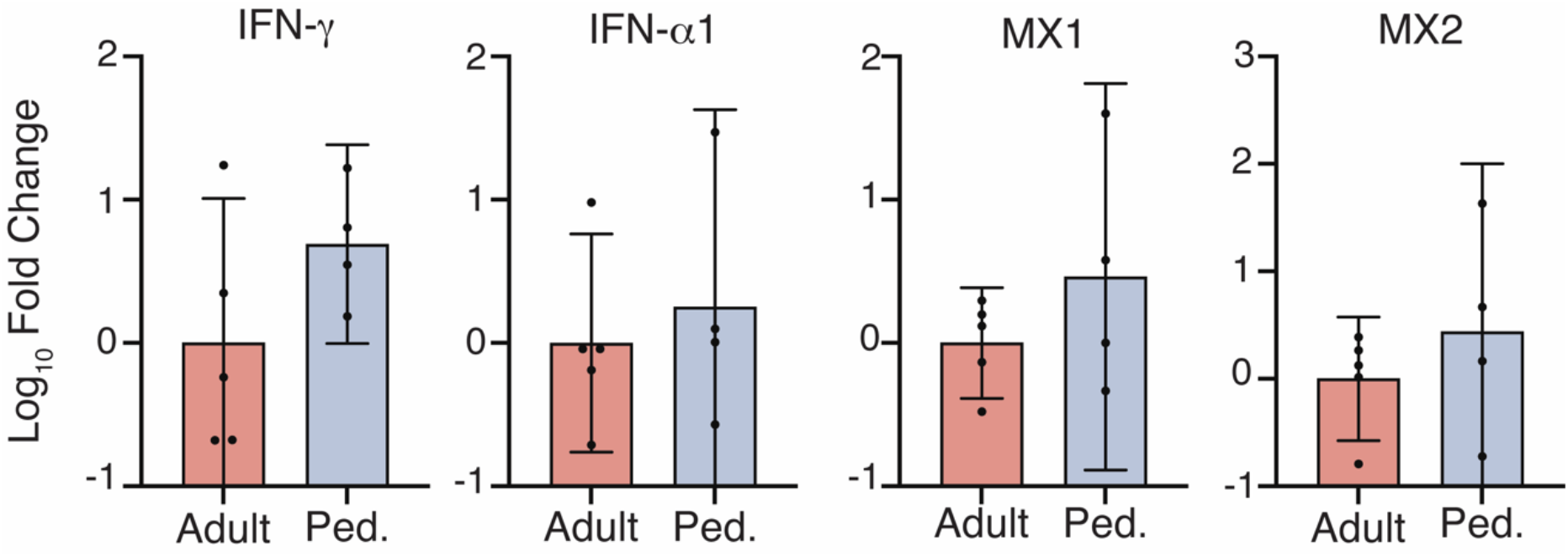
RT-qPCR of interferon-related genes. RT-qPCR was performed on indicated genes using 5 adult and 4 pediatric samples not included in the RNAseq analysis. Bars show mean ± 95% CI. Ped. = Pediatric. Fold change was calculated by the 2^CT(ref)-CT^ method using the mean adult value as the reference.

**Supplementary Table 1:** Demographic, clinical and laboratory findings of pediatric and adult patients with COVID-19.

|  | Pediatric (n=12) | Adult (n=27) | p-value* |
| --- | --- | --- | --- |
| Age, years (mean ± SD) | 6.54 ± 5.16 | 53.30 ± 22.73 | < 0.001 |
| Male:Female (n) | 5:7 | 14:13 | NS |
| Race/Ethnicity (n) |  |  |  |
| Hispanic | 6 | 18 | NS |
| Black | 3 | 6 |  |
| White | 3 | 2 |  |
| Asian | 0 | 1 |  |
| Symptom days prior to presentation | 3.75 ± 3.91 | 4.44 ± 2.84 | NS |
| Admission to hospital | 5/12 (42%) | 22/27 (81%) | 0.02 |
| Labs at admission |  |  |  |
| Hemoglobin (g/dl) | 12.60 ± 1.98 (n=5) | 12.67 ± 1.60 (n=22) | NS |
| Platelets (x1000) (cells/μl) | 330 ± 162.4 (n=5) | 253.4 ± 83.87 (n=22) | NS |
| Total white blood cell | 9500 ± 5317 (n=5) | 9682 ± 3586 (n=22) | NS |
| ALC (cells/μl) | 1680 ± 1047 (n=5) | 1550 ± 1036 (n=22) | NS |
| ANC (cells/μl) | 6760 ± 4368 (n=5) | 7291 ± 3643 (n=22) | NS |
| C-reactive protein (mg/dl) | 6.03 ± 10.48 (n=3) | 10.48 ± 7.19 (n=20) | <0.0001 |
| D-dimer (μg/ml) | 0.81 ± 0.59 (n=3) | 2.38 ± 3.42 (n=21) | <0.0001 |
| Respiratory support (n) |  |  | 0.03 |
| Room air | 12 | 16 |  |
| Supplemental oxygen | 0 | 7 |  |
| Mechanical ventilation | 0 | 4 |  |
| Length of stay (days, admitted) | 3.0 ± 1.73 (n=5) | 10.36 ± 10.85 (n=22) | <0.0001 |
| Mortality (n) | 0/12 | 4/27 | NS |

SD, standard deviation; n, number; NS, not significant; ALC, absolute lymphocyte count; ANC, absolute neutrophil count
\*Continuous variables were compared by Student's t test; categorical variables were compared by Fisher's exact test.

**Supplementary Table 2:**
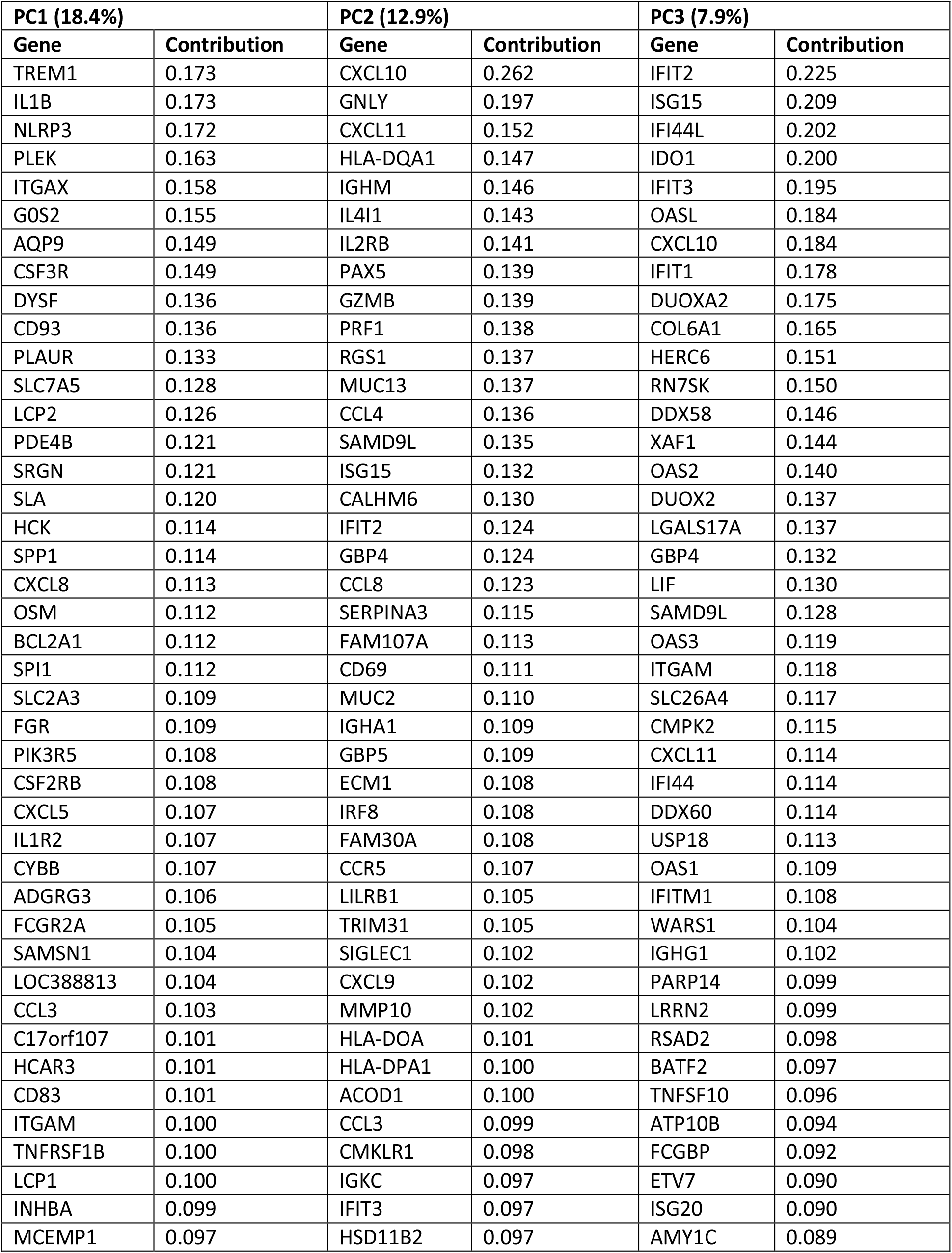
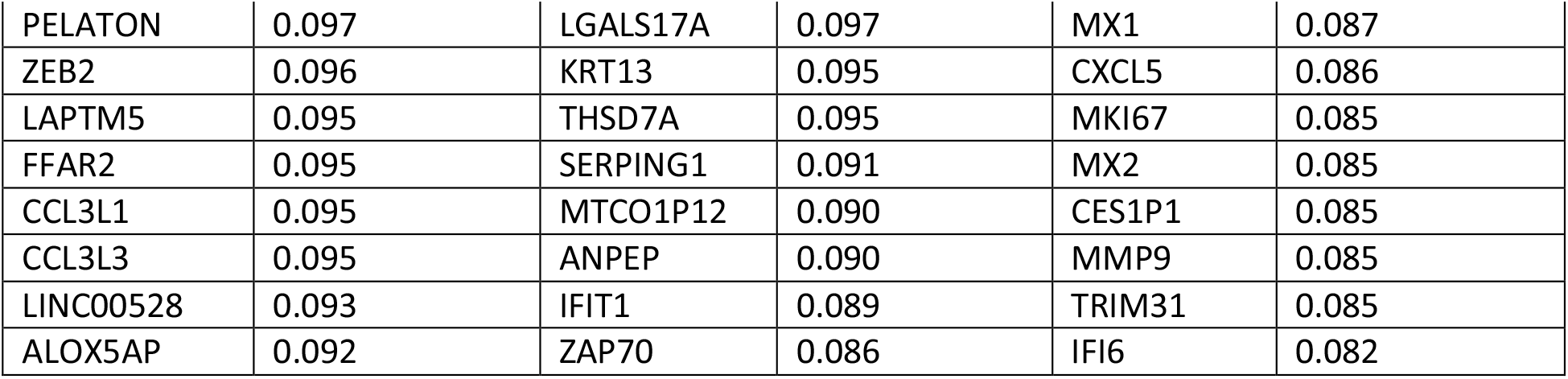
50 top contributing genes to Principal Components (PC) PC1, PC2 and PC3 and their relative contribution.

| **PC1 (18.4%)** |  | **PC2 (12.9%)** |  | **PC3 (7.9%)** |  |
| --- | --- | --- | --- | --- | --- |
| **Gene** | **Contribution** | **Gene** | **Contribution** | **Gene** | **Contribution** |
| TREM1 | 0.173 | CXCL10 | 0.262 | IFIT2 | 0.225 |
| IL1B | 0.173 | GNLY | 0.197 | ISG15 | 0.209 |
| NLRP3 | 0.172 | CXCL11 | 0.152 | IFI44L | 0.202 |
| PLEK | 0.163 | HLA-DQA1 | 0.147 | IDO1 | 0.200 |
| ITGAX | 0.158 | IGHM | 0.146 | IFIT3 | 0.195 |
| G0S2 | 0.155 | IL4I1 | 0.143 | OASL | 0.184 |
| AQP9 | 0.149 | IL2RB | 0.141 | CXCL10 | 0.184 |
| CSF3R | 0.149 | PAX5 | 0.139 | IFIT1 | 0.178 |
| DYSF | 0.136 | GZMB | 0.139 | DUOXA2 | 0.175 |
| CD93 | 0.136 | PRF1 | 0.138 | COL6A1 | 0.165 |
| PLAUR | 0.133 | RGS1 | 0.137 | HERC6 | 0.151 |
| SLC7A5 | 0.128 | MUC13 | 0.137 | RN7SK | 0.150 |
| LCP2 | 0.126 | CCL4 | 0.136 | DDX58 | 0.146 |
| PDE4B | 0.121 | SAMD9L | 0.135 | XAF1 | 0.144 |
| SRGN | 0.121 | ISG15 | 0.132 | OAS2 | 0.140 |
| SLA | 0.120 | CALHM6 | 0.130 | DUOX2 | 0.137 |
| HCK | 0.114 | IFIT2 | 0.124 | LGALS17A | 0.137 |
| SPP1 | 0.114 | GBP4 | 0.124 | GBP4 | 0.132 |
| CXCL8 | 0.113 | CCL8 | 0.123 | LIF | 0.130 |
| OSM | 0.112 | SERPINA3 | 0.115 | SAMD9L | 0.128 |
| BCL2A1 | 0.112 | FAM107A | 0.113 | OAS3 | 0.119 |
| SPI1 | 0.112 | CD69 | 0.111 | ITGAM | 0.118 |
| SLC2A3 | 0.109 | MUC2 | 0.110 | SLC26A4 | 0.117 |
| FGR | 0.109 | IGHA1 | 0.109 | CMPK2 | 0.115 |
| PIK3R5 | 0.108 | GBP5 | 0.109 | CXCL11 | 0.114 |
| CSF2RB | 0.108 | ECM1 | 0.108 | IFI44 | 0.114 |
| CXCL5 | 0.107 | IRF8 | 0.108 | DDX60 | 0.114 |
| IL1R2 | 0.107 | FAM30A | 0.108 | USP18 | 0.113 |
| CYBB | 0.107 | CCR5 | 0.107 | OAS1 | 0.109 |
| ADGRG3 | 0.106 | LILRB1 | 0.105 | IFITM1 | 0.108 |
| FCGR2A | 0.105 | TRIM31 | 0.105 | WARS1 | 0.104 |
| SAMSN1 | 0.104 | SIGLEC1 | 0.102 | IGHG1 | 0.102 |
| LOC388813 | 0.104 | CXCL9 | 0.102 | PARP14 | 0.099 |
| CCL3 | 0.103 | MMP10 | 0.102 | LRRN2 | 0.099 |
| C17orf107 | 0.101 | HLA-DOA | 0.101 | RSAD2 | 0.098 |
| HCAR3 | 0.101 | HLA-DPA1 | 0.100 | BATF2 | 0.097 |
| CD83 | 0.101 | ACOD1 | 0.100 | TNFSF10 | 0.096 |
| ITGAM | 0.100 | CCL3 | 0.099 | ATP10B | 0.094 |
| TNFRSF1B | 0.100 | CMKLR1 | 0.098 | FCGBP | 0.092 |
| LCP1 | 0.100 | IGKC | 0.097 | ETV7 | 0.090 |
| INHBA | 0.099 | IFIT3 | 0.097 | ISG20 | 0.090 |
| MCEMP1 | 0.097 | HSD11B2 | 0.097 | AMY1C | 0.089 |
| PELATON | 0.097 | LGALS17A | 0.097 | MX1 | 0.087 |
| ZEB2 | 0.096 | KRT13 | 0.095 | CXCL5 | 0.086 |
| LAPTM5 | 0.095 | THSD7A | 0.095 | MKI67 | 0.085 |
| FFAR2 | 0.095 | SERPING1 | 0.091 | MX2 | 0.085 |
| CCL3L1 | 0.095 | MTCO1P12 | 0.090 | CES1P1 | 0.085 |
| CCL3L3 | 0.095 | ANPEP | 0.090 | MMP9 | 0.085 |
| LINC00528 | 0.093 | IFIT1 | 0.089 | TRIM31 | 0.085 |
| ALOX5AP | 0.092 | ZAP70 | 0.086 | IFI6 | 0.082 |

## Supplemental Methods

### RNA sequencing

Libraries were prepared at the Yale Center for Genome Analysis using the NEBNext rRNA Depletion Kit (E6310L). Individual samples or pools from 2 patients of similar age and outcome (n=4 pools, adults) were normalized to 1.2nM and loaded on an Illumina NovaSeq S4 flow cell to generate 30M read-pairs per sample. Samples were checked for read quality and adapter contamination using FastQC and aligned to transcripts using the GENCODE transcript sequences (v33) as the reference file with Salmon (29). All analyses in R were performed using R version 4.0.3. Transcripts were mapped to genes using tximport. Differential gene expression analysis was performed with DESeq2 (30). Heatmaps were generated using the pheatmap package. Principal components analysis was performed with the *prcomp* functionusing all genes with a non-zero total read count. Prior to PCA, data were transformed with the *vst* function in DESeq2. PCA results were visualized with the factoextra package. For gene set enrichment analyses, Hallmark (h) and GO (c5.go) datasets were downloaded from MSigDB (Broad Institute) and analysis performed in R with the fgsea package using 1000 permutations.

### Real-Time Quantitative Reverse Transcription PCR

qPCR was performed using the TaqMan Gene Expression Master Mix (Applied Biosystems, 4369016). Data were analyzed by the 2^-ΔΔCt^ method, using the mean adult values as the reference. Primers/probes were from ThermoFisher: MX1 (Hs00895608_m1), MX2 (Hs01550811_m1), IFNA1 (Hs00256882_s1), IFI44 (Hs00951349_m1), IFIT1 (Hs03027069_s1), IL17A (Hs00174383_m1), IFNG (Hs00989291_m1), RPLPO (Catalog No. 4326314E).

### Cytokine and Antibody Measurements

Data from Luminex assays were acquired on a Luminex Magpix (Luminex Corporation) and analyzed in the Milliplex Analyst program (Millipore). Total IgA was measured using the IgA Human ELISA Kit (Invitrogen, BMS2096), and total IgG measured using the IgG Human ELISA Kit (Invitrogen, BMS2091). ELISA data were acquired on a Spectra Max M5 using SoftMax Pro 7.1 GxP software (both Molecular Devices).

